# MUC17 mutations and methylation are associated with poor prognosis in adult-type diffuse glioma patients

**DOI:** 10.1101/2023.01.30.23285200

**Authors:** Gabriel Cardoso Machado, Valéria Pereira Ferrer

## Abstract

Diffuse gliomas are tumors that arise from glial or glial progenitor cells. They are currently classified as astrocytoma isocitrate dehydrogenase (IDH)-mutant or oligodendroglioma IDH-mutant, and 1p/19q-codeleted, both slower-growing tumors, or glioblastoma (GBM), a more aggressive tumor. Despite advances in the diagnosis and treatment of gliomas, the median survival time after diagnosis of GBM remains low, approximately 15 months, with a 5-year overall survival rate of only 6.8%. Therefore, new biomarkers that could support the earlier diagnosis and prognosis of these tumors would be of great value. MUC17, a membrane-bound mucin, has been identified as a potential biomarker for several tumors. However, the role of this mucin in adult gliomas has not yet been explored. Here, we show for the first time, in a retrospective study and by *in silico* analysis that MUC17 is one of the relevant mutant genes in adult gliomas. Moreover, that an increase in MUC17 methylation correlates with an increase in glioma malignancy grade. Patients with MUC17 mutations had a poorer prognosis than their wild-type counterparts in both GBM and non-GBM glioma cohorts. We also analyzed mutational profiles that correlated strongly with poor survival. Therefore, in this study, we present a new potential biomarker for further investigation, especially for the prognosis of adult diffuse gliomas.

## 1 Introduction

Glial tumors are the most common primary malignant brain tumors of the central nervous system (CNS)[1]. Diffuse gliomas represent a heterogeneous group of brain tumors that arise from neuroglial stem/progenitor-like cells, and exhibit genetic and epigenetic malignant modifications[2].

Glioblastoma (GBM) is the most common CNS malignancy, accounting for 49.1% of all primary CNS malignancies[1]. GBM is characterized by a poor prognosis and limited therapeutic options[3]. It has a median survival time of 15 months after diagnosis and a 5-year overall survival (OS) rate of only 6.8%, which varies by age and sex[4]. This tumor is 1.6 times more common in men[1] and has a median age at diagnosis of 64 years, with a peak at 75-84 years[5]. Despite the standard treatment of GBM consisting of surgery, radiotherapy, chemotherapy[6], and advances in brain tumor therapies[7,8], the disease remains incurable. Moreover, it shows a high rate of treatment resistance and recurrence[4,9].

Since 2016, the molecular signatures of genes associated with the diagnosis and prognosis of diffuse gliomas have been included in the World Health Organization (WHO) classification system[10]. Among the key markers, mutations in IDH genes and co-deletions of chromosome arms 1p and 19q were the first molecular criteria used to classify astrocytoma’s and oligodendrogliomas, respectively[11,12]. In general, these deletions and mutations predict a better clinical picture; therefore, they are rare in GBM and common in less aggressive gliomas[13,14]. Methylation status of the O^6^-methylguanine-DNA methyltransferase (MGMT) promoter region is another important biomarker of diffuse gliomas[15]. Epigenetic inactivation of MGMT is associated with increased survival in patients with glioma, and predicts benefit in using alkylating agents for chemotherapy in patients with GBM[16,17].

According to the WHO glioma classification, updated in 2021, tumor grade (from 2 to 4) reflects a combination of histological features, but with well-defined genetic alterations. In this new classification, gliomas are simply grouped into GBM IDH-wildtype, which is thus the most aggressive form of diffuse glioma, while astrocytoma IDH-mutant, oligodendroglioma IDH-mutant, and 1p/19q-codeleted comprise less aggressive tumors. We refer to these as non-GBM glioma tumors[18,19].

The latest WHO classification of gliomas underlines the importance of molecular knowledge for the diagnosis and prognosis of gliomas.

Mucins are a family of high-molecular-weight glycoproteins, encoded by twenty-one currently known genes in the human genome[20]. Based on structure and cellular localization, this family is divided into two main groups: secreted and membrane-anchored mucins[21]. Membrane-bound mucins share a single-pass transmembrane domain and are composed of at least ten mucin types, including MUC17.

MUC17 is located at locus 7q22.1, which[22] encodes the third largest membrane mucin (4493 amino acids), whose PTS domain (proline/threonine/serine) occupies 4073 amino acids with more than 1,600 O-glycosylation sites[23–25]. MUC17 has been identified as a potential biomarker in several tumors, including breast cancer[26–28], gastric cancer[29–31], colon cancer[32–34], bile duct cancer[35], laryngeal squamous cell carcinoma,[36] and pancreatic ductal adenocarcinoma[37]. However, little is known about the clinical association between MUC17 molecular changes and gliomas. To date, only one study has shown that MUC17 mutations predict a favorable prognosis in pediatric-type malignant high-grade gliomas[38].

Therefore, this study is the first to describe MUC17 mutations and epigenetic modifications related to tumor grade, clinical features, and prognosis of adult diffuse gliomas. Here, we have shown that MUC17 mutations are part of the mutational burden in adult gliomas and that an increase in MUC17 methylation correlates with an increase in glioma malignancy grade. MUC17 mutations correlated with poor prognosis in both GBM patients and non-GBM gliomas, and we analyzed which mutational profile was more strongly associated with poor survival. Thus, we identified a new potential biomarker to be explored, particularly for the prognosis of diffuse glioma in adults.

## 2 Material and Methods

### 2.1 Glioma mutational burden analysis

Data on the most frequently mutated genes in gliomas were collected from the Broad Institute of Massachusetts Institute of Technology (MIT) and the Harvard Portal (http://firebrowse.org/)[39]. Three significance metrics were calculated for each gene using MutSigCV[40]. These measure the significance of the mutation load. MutSigCV determines the P-value for observing a given number of non-silent mutations in the gene, given the background model determined by silent (and non-coding) mutations in the same gene and neighboring genes of the covariate space. The following contextual categories were used: transitions at CpG dinucleotides, transitions at other C-G base pairs, transversions at C-G base pairs, mutations at A-T base pairs, and indels. Therefore, in this analysis, we selected significantly mutated genes in adult-type diffuse gliomas for comparison with normal samples.

### 2.2 MUC17 methylation profile in gliomas

The clinical and methylation data of 14.776 genes from 159 patients (8 normal and 151 glioma patients) were downloaded from the Chinese Glioma Genome Atlas (CGGA) (http://www.cgga.org.cn/)[41]. MUC17 methylation parameters from the 151 patients with gliomas (containing both primary and recurrent gliomas) were assessed and scored from 0 (hypomethylated) to 1 (hypermethylated)[42]. We divided the cohort into GBM and non-GBM patients according to different malignancy grades (2-4), sex, and age (over or under 40 years). For statistical analysis, we first applied the normality and log normality tests (D′Agostino and Pearson tests). We verified that the data did not follow a normal distribution. Therefore, we used the Mann-Whitney test for the comparison of the two groups and the Kruskal-Wallis test for comparison of three or more groups, using Dunńs multiple comparisons test when applicable. Methylation graphs were generated using GraphPad software version 9.

### 2.3 Data for comparative analysis between GBM and non-GBM samples

Using cBioPortal, we selected the following studies to compose the GBM cohort:141 samples from two multiomics studies[43,44], 783 samples from two TCGA studies[45,46], 619 samples from TCGA - GDAC Firehose Legacy (https://gdac.broadinstitute.org/runs/stddata2016_01_28/data/GBM/20160128/), and 592 samples from TCGA – PanCancer Atlas[47–56]. After selecting the studies, we restricted the classification for further analysis: 634 samples from glioblastoma (29.7%) and 206 samples from glioblastoma multiforme (9.6%). We excluded the specimens classified as “glioma” (N= 1295; 29,75%). Therefore, we analyzed 840 samples from 833 patients with GBM.

For the non-GBM cohort, the selected studies used were: 530 samples from The Cancer Genome Atlas (TCGA) - GDAC Firehose Legacy (https://gdac.broadinstitute.org/runs/stddata2016_01_28/data/LGG/20160128/); 514 samples from the TCGA – PanCancer Atlas[47–56]; 444 samples from The Glioma Longitudinal AnalySiS (GLASS) consortium[57]; 1095 samples sequenced from the MSK-IMPACT platform in two different studies[58,59]; 61 samples sequenced by exome analysis in a study on LGG evolution[60]; and 1122 samples sequenced by TCGA[61]. After study selection, we refined the search for further analysis: 1167 samples of diffuse glioma (31%); 447 specimens of oligodendroglioma (11.9%); 341 samples of anaplastic astrocytoma (9.1%); 282 specimens of astrocytoma (7.5%); 280 samples of oligoastrocytoma (7.4%); 178 specimens of diffuse astrocytoma (4.7%); 78 samples of anaplastic oligoastrocytoma (2.1%), and 71 specimens of anaplastic oligodendroglioma (1.9%). We excluded histology with <1% incidence, comprising a total of 32 specimens. Therefore, we analyzed 2884 samples from 2691 non-GBM patients.

### 2.4 Overall survival analysis

Survival data revealed the last known day up to which the patient survived. Survival comparisons between the selected genes were performed using the Kaplan-Meier method and the log-rank (Mantel-Cox) statistical test using GraphPad Prism (version9). The median survival time was calculated as the shortest survival time for which the survival function was equal to or less than 50%. Overall survival (OS) data were obtained using cBioPortal as described previously. First, we compared patients with wild-type MUC17 with those with mutated MUC17 among the 2884 non-GBM glioma patients and 840 GBM patients.

We then compared patients with MUC17 mutations with those with clinically important, and commonly mutated genes in gliomas. For this analysis, data from 593 non-GBM and 193 GBM samples were used. The mean survival time was calculated for non-GBM glioma samples based on mutations in MUC17, ATRX chromatin remodeler, telomerase reverse transcriptase (TERT), tumor protein p53 (TP53), and isocitrate dehydrogenase 1 (IDH1). For GBM, OS was estimated based on mutations in MUC17, phosphatidylinositol-4,5-bisphosphate-3-kinase catalytic subunit alpha (PIK3CA), phosphatase and tensin homologue (PTEN), TP53, and epidermal growth factor receptor (EGFR). In this analysis, only samples that exclusively contained the mentioned mutations were included and overlapping mutation samples were excluded.

We also analyzed these patients by classifying them according to sex (male and female) and age (under and ≥ 40 years). Additionally, we selected the clinical data of 21 patients to analyze the following parameters: GBM or non-GBM subtype, grade, radiotherapy status, and chemotherapy status.

### 2.5 IDH1 status

Clinical data from patients with glioma were obtained from the cBioPortal (https://www.cbioportal.org/)[62,63]. IDH mutation status was analyzed in 1130 non-GBM samples with clinically important mutations (ATRX, IDH1, TERT, and TP53) and MUC17 mutations. For analysis of the 146 GBM samples, these status data were unavailable for comparison. A percentage bar chart was created using cBioPortal.

### 2.6 MUC17 mutation profile

The glioma mutation profile was generated at the Broad Institute of Massachusetts Institute of Technology (MIT) and the Harvard Portal (http://firebrowse.org/viewGene.html) using the iCoMut Beta. Glioma specimens (1115) were profiled and classified into synonymous in-frame indels, other non-synonymous mutations, missense mutations, splice sites, frameshift mutations, and nonsense mutations.

The MUC17 mutational profile of patients with GBM was created using data from the Tumor Portal (http://www.tumorportal.org/view?geneSymbol=MUC17). The mutational profiles of 29 GBM samples with MUC17 mutations were analyzed. Mutations were classified as synonymous or missense, in-frame insertions or deletions, frameshift insertions or deletions, and nonsense or splice-site mutations. Blue, white, and red bars are the log2 distributions of the somatic copy number alteration ratio (–1 to +1)[64]. For missense mutations, we analyzed the frequency of base-pair changes and median OS.

## 3 Results

### 3.1 Mutated MUC17 is a significant gene in gliomas

MUC17 has a 6% mutation frequency in gliomas and is one of the significantly mutated genes in these patients. It reaches the eighth position, preceded only by IDH1, TP53, ATRX, EGFR, PTEN, CIC, and PIK3CA genes (Figure 1A). In gliomas, most mutations in MUC17 are missense mutations, but there are a minority of synonymous mutations (Figure 1A).

**Figure 1.**
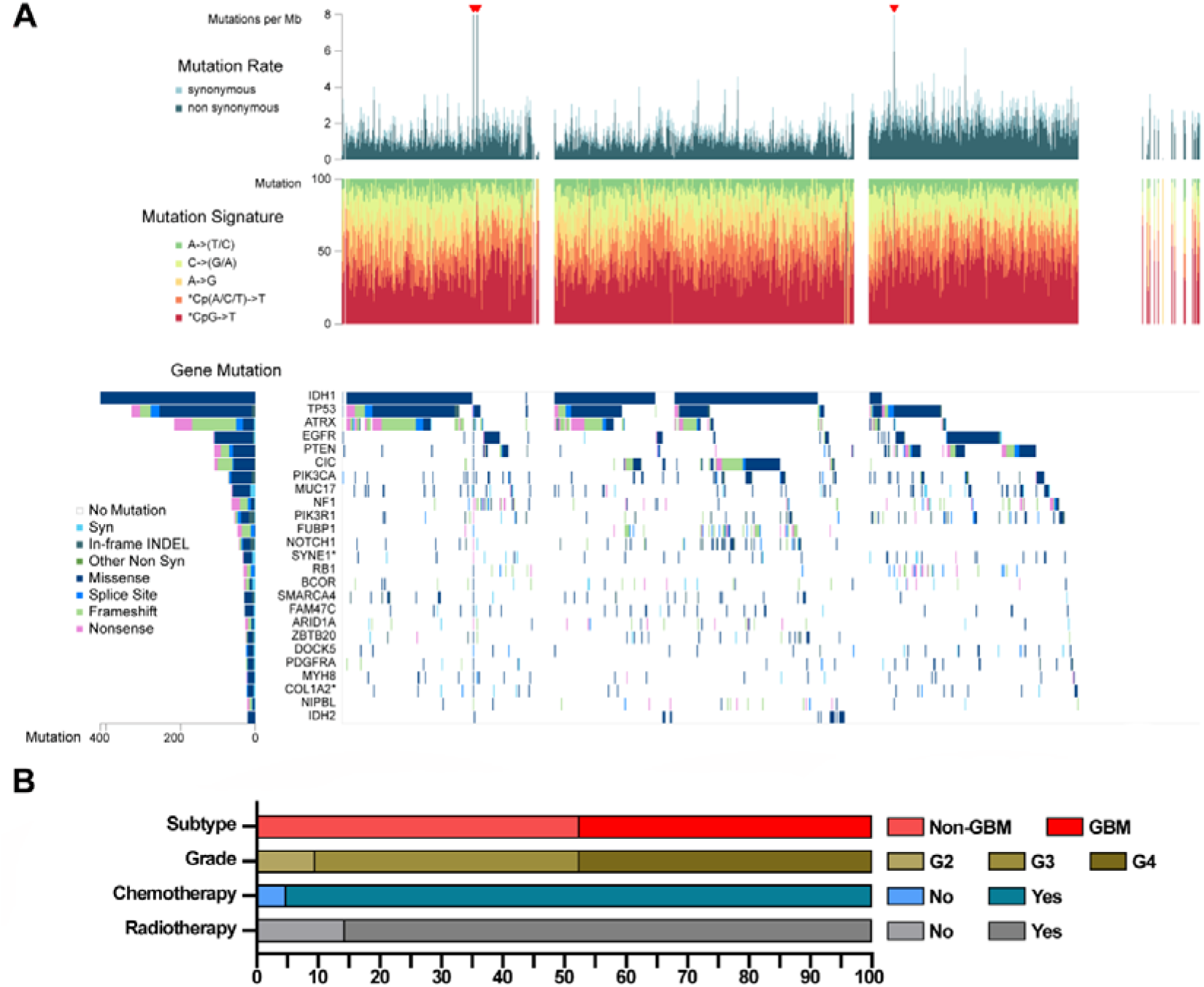
MUC17 is among the significantly mutated genes in glioma patients. **(A)** The matrix represents individual mutations in patient samples, color-coded by type of mutation, for the significantly mutated genes. The barplot on the left of the matrix shows the number of mutations in each gene. MUC17 is the eighth most mutated gene in adult glioma patients and contains the most missense mutations and a minority of synonymous mutations **(B)** Clinical profile of glioma patients with MUC17 mutations from the cBioPortal cohort. The stacked bar chart illustrates the distribution of patients by subtype (non-GBM or GBM), grade (2, 3 and 4), radiotherapy and chemotherapy status (yes= received treatment or no= treatment not received). Most MUC17-mutated patients are classified as non-GBM, WHO grade 4, and received chemotherapy and radiotherapy treatment.

Of the patients with MUC17 mutations, 47.62 % were classified as WHO grade 4, 42.86 % as grade 3, and 9.52 % as grade 2 (Figure 1B). Of these, 47.62% were classified as GBMs and 52.38% as non-GBMs. When we analyzed these patients by treatment status, 85.71% of the patients were treated with radiotherapy and 95.24% required chemotherapeutics.

### 3.2 MUC17 gene methylation is correlated to glioma staging and increased in GBM patients

To assess the methylation pattern of MUC17 in gliomas, we utilized clinical and methylation data from the CGGA. Between the two subtype groups, we observed a significant increase (p =0.0015) in MUC17 methylation in GBM (median=0.81, q1=0.75, q3=0.85, N=43) compared with non-GBM patients (median=0.71, q1=0.62, q3= 0.83, N=108) (Figure 2A). We also found differences in MUC17 methylation among tumor grades 2, 3, and 4 (p = 0.0066). Specifically, we found a significant increase in MUC17 methylation in patients with grade 4 tumors (median= 0.81, q1=0.75, q3=0.85, N=43) compared with that in patients with grade 2 (median=0.71, q1=0.58, q3=0.87, N=61; p=0.0217) and those with grade 3 tumors (median= 0.70, q1=0.62, q3=0.82, N=47; p=0.0116) (Figure 2B). No significant difference was found in MUC17 methylation between patients with grade 2 and 3 tumors (p > 0.9999).

**Figure 2.**
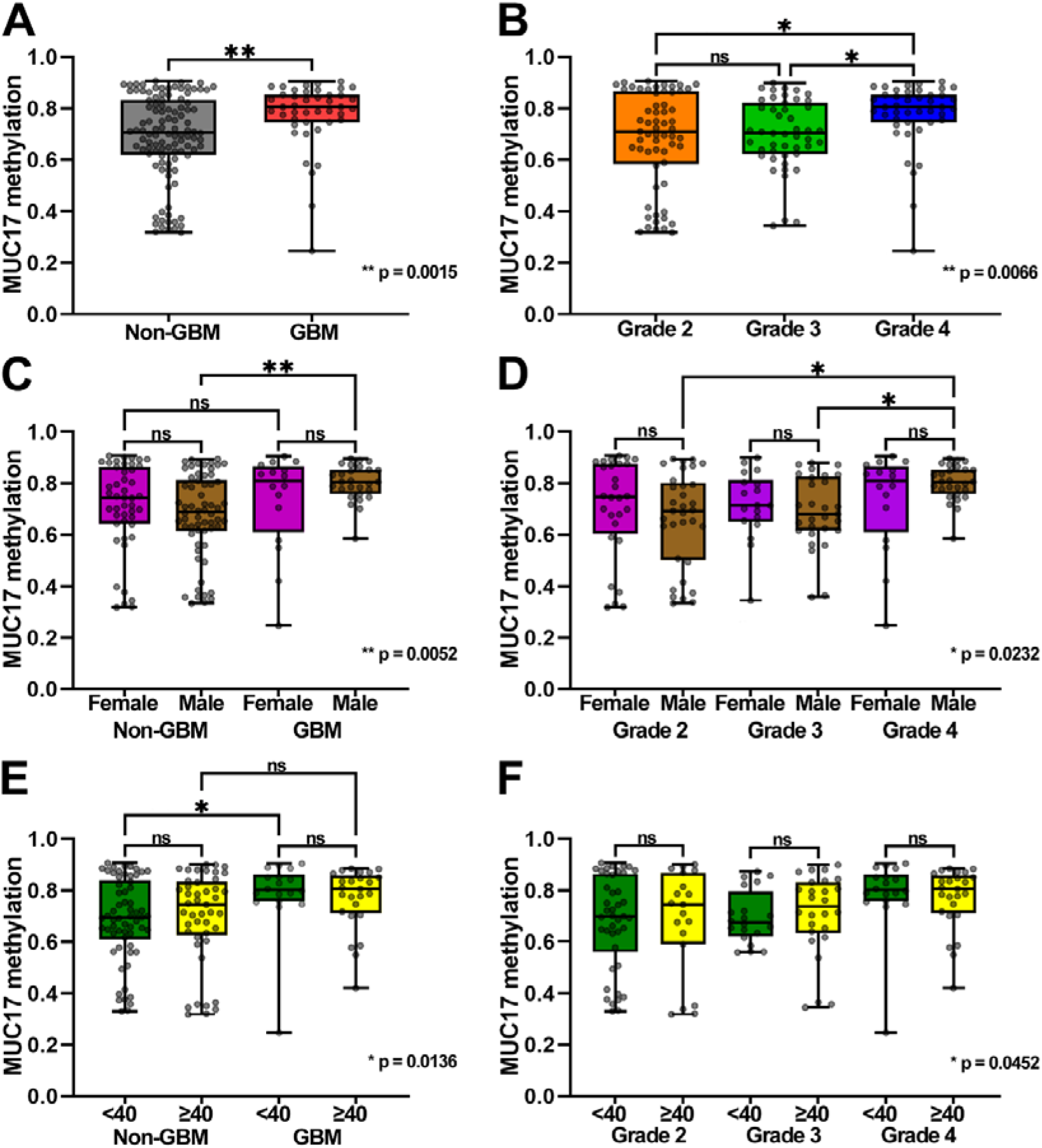
MUC17 methylation is increased in grade IV and GBM gliomas. (A) Methylation of the MUC17 gene comparing the pattern of adult-type diffuse gliomas: GBM and non-GBM groups. The GBM group shows a significant increase in MUC17 methylation (**p=0.0015) compared with non-GBM patients. (B) MUC17 methylation among samples of different glioma grades. Grade 4 samples exhibit the highest level of MUC17 methylation compared with lower grades (**p=0.0066). Grade 4 samples show significantly higher methylation than grade 3 (*p= 0.0116) and grade 2 (*p= 0.0217) samples. (C) MUC17 methylation in glioma subtypes and sexes. There is a difference in the methylation pattern of GBM and non-GBM groups when we subdivide them into male and female (**p=0.0052). Interesting, males from the GBM group have higher methylation than non-GBM counterparts (*p= 0.0024). (D) Methylation of MUC17 gene among different grades and sexes. There is a difference in the methylation pattern among grades when we subdivide them into male and female (*p=0.0232). Additionally, grade 4 males have higher methylation patterns than patients with grade 2 (*p= 0.0188) and grade 3 tumors (*p= 0.0312). Under and above 40 years-old patients according to glioma subtypes (E) and grades (F). There are differences in GBM and non-GBM groups (*p=0.0136) and grades (*p=0.0452) depending on age. Younger GBM patients show a higher methylation pattern than younger non-GBM patients (*p= 0.0193). There are no significant differences when comparing younger and older patients within the same grade, at the same age, with different grades.

Next, we investigated the methylation profile of MUC17 considering the sex of patients with glioma in the GBM and non-GBM groups. A significant difference (p= 0.0052) was observed between the groups (Figure 2C). Interestingly, when we compared patients of the same sex, but with different glioma subtypes individually, we found a significant increase (p= 0.0024) in MUC17 methylation in men with GBM (median=0.81, q1= 0.76, q3=0.85, N=27) compared with non-GBM male patients (median= 0.69, q1= 0.61, q3= 0.81, N=62). There was no significant difference (p >0.9999) in the methylation patterns between female patients with GBM (median= 0.81, q1=0.61, q3=0.87, N=16) and non-GBM female patients (median=0.74, q1= 0.64, q3=0.86, N=46).

When we stratified the glioma patients by grade and subdivided them according to sex, we found a significant difference in MUC17 methylation among the groups (p= 0.0232) (Figure 2D). When we paired patients of the same sex along the tumor grades, we found a significant increase in MUC17 methylation in male patients with grade 4 tumors (median= 0.81, q1= 0.76, q3= 0.85, N=27) compared with those with grade 3 (median= 0.68, q1= 0.62, q3= 0.83, N=29; p= 0.0312) or grade 2 tumors (median= 0.69, q1= 0.50, q3= 0.80, N=33; p= 0.0188). However, no statistically significant differences in MUC17 methylation were observed between female patients with grade 4 (median= 0.81, q1= 0.61, q3= 0.87, N=16), grade 3 (median= 0.71, q1= 0.65, q3= 0.81, N= 18; p >0.9999), or grade 2 tumors (median= 0.75, q1= 0.60, q3= 0.88, N=28; p >0.9999).

Furthermore, we assessed the methylation profile of MUC17 in patients with GBM and non-GBM or in different grades, segregating them according to age (below and above 40 years). There was a significant difference (p= 0.0136) in MUC17 methylation between the age groups (Figure 2E). When we paired patients within the same age group in the two glioma subtypes, we found a significant increase in MUC17 methylation (p= 0.0193) in younger patients with GBM (median = 0.80, q1 = 0.76, q3 = 0.86, N = 18) than in younger patients without GBM (median= 0.69, q1=0.61, q3= 0.84, N= 62) (Figure 2E). There was no significant difference between older patients with GBM (median = 0.81, q1 = 0.71, q3 = 0.86, N = 25) and older patients without GBM (median= 0.74, q1= 0.63, q3=0.83, N=45) (p= 0.4955).

We found a significant difference (p=0.0452) in MUC17 methylation across different tumor grades according to age (Figure 2F). We observed the followed methylation results: grade 4 patients <40 years old (median= 0.80, q1= 0.76, q3= 0.86, N=18) and ≥ 40 years old (median= 0.81, q1= 0.71; q3= 0.86, N=25); grade 3 patients <40 years old (median= 0.67, q1= 0.62, q3= 0.80, N=20) and ≥ 40, (median= 0.74; q1= 0.63, q3= 0.83, N=26); grade 2 patients <40 years old (median= 0.70, q1=0.56; q3= 0.86, N=42) and ≥ 40 years old (median= 0.74, q1= 0.59, q3= 0.87, N= 19). No statistically significant differences were observed between younger and older patients with the same tumor grade or among the same age group (non-significance bars not shown).

### 3.3 Patients with MUC17 mutation have poorer OS rates than wild-type counterparts in GBM and non-GBM tumors

When we analyzed a larger cohort of patients and divided them into non-GBM (Figure 3A) and GBM samples (Figure 3B), we found that patients with MUC17 mutations in both non-GBM gliomas (p=0.0001) and GBM (p = 0.0369) had a worse prognosis than patients with MUC17 wild-type groups. In the non-GBM glioma cohort, median survival was 39.0 months in mutant patients versus 70.2 months in wild-type patients; and 13.9 versus 14.46 months in GBM patients, respectively.

**Figure 3.**
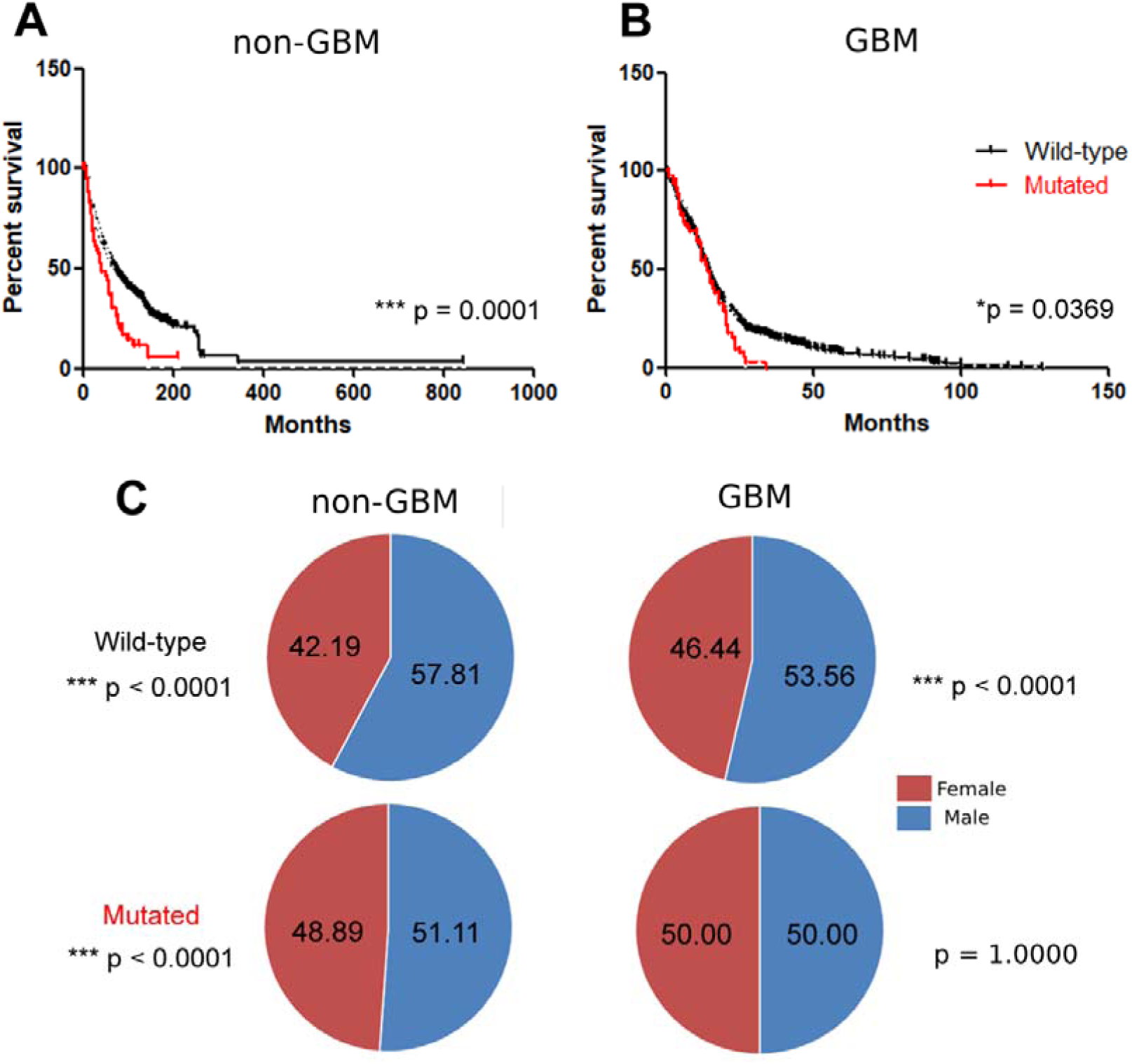
Adult diffuse glioma patients with MUC17 mutation have worse prognosis than wild-type counterparts. Kaplan-Meier OS curve of (A) non-GBM gliomas and (B) GBM patients. Patients with mutated MUC17 genes have worse prognosis than their wild-type counterparts in both the non-GBM (***p = 0.001) and GBM (*p = 0.0369) cohorts. (C) Gender distribution plots in non-GBM and GBM patients with wild-type or mutated MUC17. Most patients without MUC17 mutation are male (57.81% in non-GBM gliomas and 53.56% in GBM). Most of non-GBM patients with MUC17 mutation are male (51.11%, p <0.0001), but in GBM patients bearing the mutation there is no difference between sexes (p = 1,0000).

To determine whether the MUC17 mutation preferentially affects a specific sex or age range, we analyzed these patients in this regard (Figure 3C). We found that among patients without MUC17 mutations, males were more affected by both non-GBM gliomas and GBM tumors (p < 0.0001). In the mutant MUC17 cohort, males were more affected by this mutation in non-GBM gliomas (p <0.0001); however, there were no sex differences in patients with GBM (p = 1.0000). We divided the non-GBM glioma cohort into adult patients under and over 40 years old and the GBM cohort into adult patients under and over 60 years old. The results showed that in non-GBM glioma patients, MUC17 mutations occurred more frequently in patients over 40 years old (58.05%, p <0.0001), while in GBM patients, MUC17 mutations occurred more frequently in patients under 60 years (51.51%, p <0.0001).

### 3.4 MUC17 mutation is clinically relevant in both GBM and non-GBM glioma cohorts

When comparing the OS of patients with MUC17 mutation to the most frequently mutated genes in non-GBM group (IDH1, TP53, ATRX and TERT), we found that patients with mutated MUC17 have the second worst OS (p = 0.0001) (Figure 4A). Their median survival was 10.60 months, behind mutant ATRX (6.74 months) and followed by mutant TP53 (15.90 months), TERT (43.50 months), and IDH1 (154.35 months). A similar analysis was performed to compare the OS of MUC17 mutant GBM patients (Figure 4B) to that of patients with mutations in clinically relevant genes in GBM (PTEN, TP53, EGFR, and PI3KCA). A worse mean OS was found in patients with mutations in EGFR (11.24 months), followed by PIK3CA (13.35 months), MUC17 (14.93 months), PTEN (16.80 months), and TP53 (19.82 months) genes (p=0.0347).

**Figure 4.**
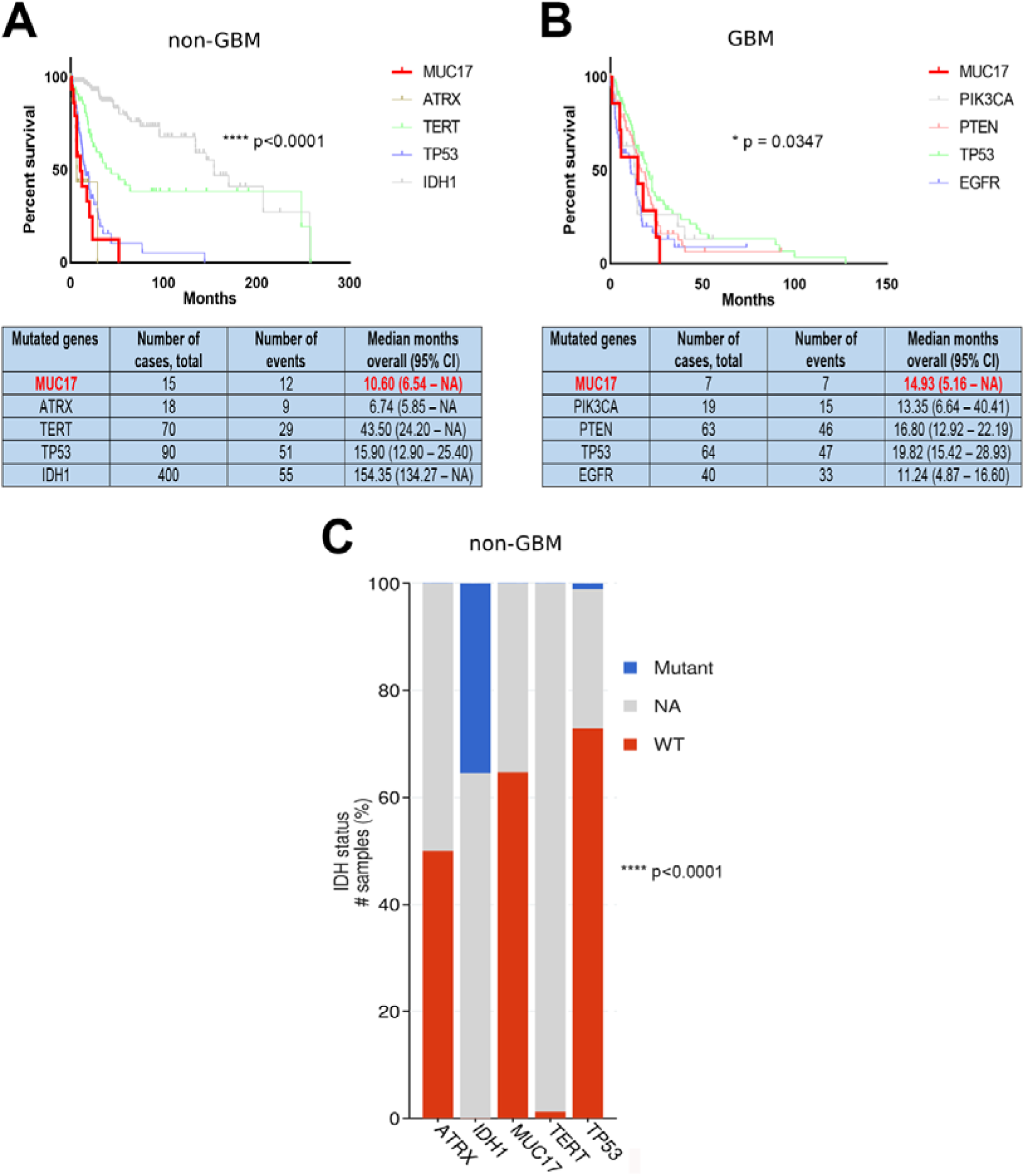
Mutations in MUC17 have clinical significance in glioma patients. (A) Analysis of the OS curve in non-GBM patients with MUC17, ATRX, TERT, TP53 and IDH1 mutations. Worse OS is observed in patients with ATRX mutations (6.74 months), followed by non-GBM patients with MUC17 (10.60 months), TP53 (15.90 months), TERT (43.50 months), and IDH1 (154.35 months; ****p<0.0001). (B) Analysis of the OS curve in GBM patients with mutations in MUC17, PIK3CA, PTEN, TP53 and EGFR. Worse OS is observed in patients with EGFR (11.24 months), followed by GBM patients with PIK3CA-(13.35 months), MUC17-(14.93 months), PTEN-(16.80 months) and TP53-mutations (19.82 months; *p=0.0347). (C) Analysis of IDH status in non-GBM glioma patients with these different gene mutations: 64.71% and 72.92% of samples from patients with MUC17 and TP53 mutations carry IDH wild-type, indicating a poor prognosis in glioma.

When we analyzed the non-GBM cohort for IDH status, we found that the percentage of wild-type IDH patients increased from mutant IDH1 patients (0%), to mutated TERT (1.23%), ATRX (50.0%), MUC17 (64.71%) and TP53 patients (72.92%).

### 3.5 C >T base change was the most abundant and represents a worse prognosis within MUC17 missense mutations in the GBM cohort

Among GBM patients with MUC17 mutations, 75.86% (22/29) had a missense mutation, 20.69% (6/29) had a silent mutation, and 3.45% (1/29) had a splice-site mutation (Figure 5A). We determined the frequency of base changes in missense mutations in this GBM cohort and calculated the median OS for each change (Figure 5B). At 40.91%, the base change C >T was the most abundant type of missense mutation, and this was the base change with the worst OS rate (5.16 months). The prognosis increased as follows: C >T (40.91%), OS 5.16 months; G >A (18.18%), OS 7.265 months; A >G (18.18%), OS 12.165; T >C (4.55%), OS 14.93 months; C >G (13.64%), OS 19.66 months; G >C (4.55%), OS 20.38.

**Figure 5.**
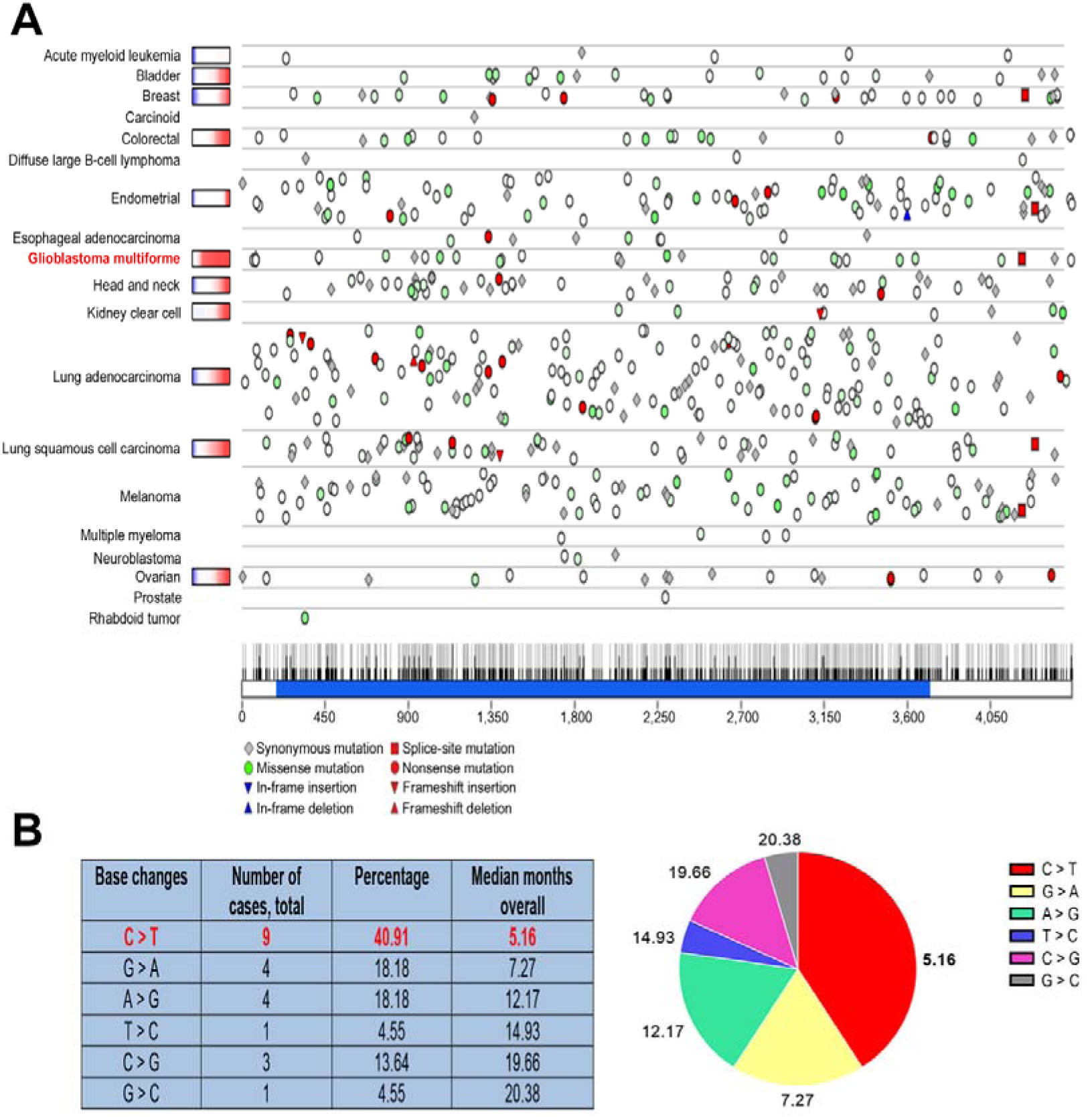
Profile of MUC17 mutations in GBM patients. (A) Graph showing the types of MUC17 mutations in GBM cohorts (N=66). Missense green saturation indicates evolutionary conservation of the mutant positions. Blue-white-red bars are log2 distributions of somatic copy number alterations ratio (1 to +1). (B) Table and graph showing the frequency of base changes (single nucleotide polymorphism, SNP) in missense mutations in GBM. C > T is the most common base change (40.91%) and these patients have the worst mean OS (5.16 months).

In the cohort analyzed, the only conserved mutation was G >A at position 100,674.926 base pairs on chromosome 7, converting valine at amino acid position 77 to methionine. This mutation was observed in two patients with an OS of 3.29 and 2.91 months.

## 4 Discussion

According to the latest WHO classification, adult diffuse gliomas can be divided into three categories: (1) Astrocytoma IDH-mutant representing IDH-mutated tumors with intact chromosome arms 1p/19q, and often with ATRX and/or TP53 mutations. Tumors in this category can be classified as WHO 2, 3, or 4 depending on their histological and molecular features[19,65]. (2) Oligodendroglioma, IDH-mutant and 1p/19q-codeleted which are tumors with mutations in IDH that have concomitant loss of chromosome arms 1p and 19q; frequently accompanied by wild-type ATRX and TP53. These tumors are classified as WHO grade 2 or 3, based on histological and molecular findings. [19,66]. (3) Glioblastoma, IDH wild-type refers to diffusely infiltrating IDH wild-type gliomas. They exhibit at least one of the following features: microvascular proliferation, necrosis, EGFR amplification, TERT promoter mutation, and/ or simultaneous chromosome 7 gain and chromosome 10 loss (+ 7/− 10). These tumors exhibit the most aggressive behavior and the worst clinical outcomes[19,67]. As a retrospective study, based on the disponible database information, we were able to classify patients with glioma into two cohorts: the GBM group and the non-GBM group; the latter included all other glioma categories that were not GBM.

Despite advances in our understanding of the underlying pathogenesis of gliomas and advances in treatment modalities, diffuse gliomas remain surgically incurable, and the 5-year survival rate for GBM remains approximately 6.8%[65]. Therefore, new biomarkers that could support the earlier diagnosis and prognosis of these tumors have been intensively explored[68].

However, the role of MUC17 in gliomas has been poorly studied. Hu et al. (2022)[38] examined MUC17 mutations in diffuse hemispheric glioma H3 G34-mutant (G34-DHG), a new type of pediatric diffuse high-grade glioma. These authors found that MUC17 was one of the genes frequently mutated in this type of tumor and that mutated MUC17 tended to indicate a favorable prognosis. However, there is currently a consensus that the gain of chromosome 7, where the MUC17 gene is located, predicts a poor prognosis in adult gliomas[19,69–71]. To our knowledge, this is the first study exploring MUC17 mutations in adult-type diffuse gliomas.

We found that most glioma patients with MUC17 mutations were in the non-GBM group, classified as WHO grade 4, and underwent radiotherapy and chemotherapy. Therefore, we found that MUC17 mutation in adult-type gliomas generally predicts the aggressive behavior of these tumors. Marchocki et al. (2022) also found that MUC17 mutation represents a worse scenario in ovarian cancer[72]. They compared exonic non-synonymous mutations in pre-and post-neoadjuvant chemotherapy samples from the same patient with high-grade serous ovarian carcinoma. They found no trends in the mutational burden following exposure to neoadjuvant chemotherapy in platinum-resistant versus platinum-sensitive patients. Most mutated genes were unique to each case. However, four mutated genes appeared exclusively in platinum-resistant cases, and MUC17 was one of the genes observed.

In our study, we verified that MUC17 gene methylation status was increased in patients with WHO grade 4 compared with those with lower grades. Additionally, methylation of MUC17 was higher in GBM than in non-GBM groups, being especially high in male and younger patients with GBM. Therefore, an increase in gene methylation correlates with a more aggressive tumor profile. The methylation of a gene promoter usually decreases its expression[15,73]. Epigenetic regulation of MUC17 gene expression, such as promoter methylation and histone modification, in pancreatic cancer was first reported in 2011. These results indicate that DNA methylation and histone H3-K9 modification in the 5’ flanking region play crucial roles in MUC17 expression. The authors found that the hypomethylation status of MUC17 was observed in patients with pancreatic ductal adenocarcinomas and that the status of the MUC17 promoter could be a novel epigenetic marker for the diagnosis of this cancer type[37,73]. MUC17 expression is reduced in hyperplastic polyps (p=0.0003), tubular and tubulovillous adenomas (p<0.0001), and colon cancers (p<0.0001), in comparison with its high expression in the surface epithelium and crypts of the colonic mucosa[33]. Interestingly, MUC17 is downregulated in *H. pylori*-infected gastric cancer (GC) tissues and cells, and is associated with poor survival in these patients. This downregulation was attributed to DNA methyltransferase 1 (DNMT1)-mediated methylation of the MUC17 promoter and was associated with GC cell proliferation and colony formation[29].

The mutant MUC17 OS rates were worse than those of other clinically relevant genes in gliomas. Additionally, we found that GBM and non-GBM glioma patients with MUC17 mutations had a poorer prognosis than patients with MUC17 wild-type status, with a stronger effect in the non-GBM cohort. In agreement with our work, in patients with biliary tract cancer, several mutated genes were found to have negative survival effects, and one of the strongest survival effects belonged to a novel recurrent deletion at 7q22.1, which excises MUC17[35]. How specific MUC17 mutations culminate in an aggressive phenotype requires further investigation. We hypothesized that these mutations would affect resistance to therapy. *In vitro* analysis of drug susceptibility and survival analysis of expression levels in patient cohorts identified MUC17 as a mediator and predictive marker of response to chemotherapy in breast cancer[26]. Additionally, malignant proliferation of non-small cell lung cancer (NSCLC) cells induced by gefitinib/osimertinib resistance is due to hypermethylation of the MUC17-specific promoter caused by the UHRF1/DNMT1 complex, which activates the NF-κB signaling pathway[74]. As mentioned earlier, mutated MUC17 is one of four genes that have emerged exclusively in platinum-resistant cases following post-neoadjuvant chemotherapy in high-grade serous ovarian carcinoma [72]. We also found that the mutation in MUC17 changed valine at position 77 to methionine in two patients with GBM. Among the metabolic differences between normal and tumor cells, the apparent dependence of GBM tumors on exogenous methionine is a critical factor that is not well understood. Methionine links the tumor microenvironment to cellular metabolism, epigenetic regulation, and mitosis. However, further studies are required to confirm this hypothesis[75].

Overall, we describe here, for the first time that MUC17 mutations account for the mutational burden of adult-type gliomas, and that MUC17 gene methylation and mutations are associated with poor prognosis in both non-GBM gliomas and GBM cohorts. Further studies are required to verify the roles of these mutations and hypermethylation in the pathobiology of gliomas in adults. However, we have opened a new avenue to explore potential biomarker that may assist in the diagnosis and prognosis of adult patients with diffuse glioma.

## Data Availability

All data produced in the present study are available upon reasonable request to the authors

## Acknowledgements

The authors thank all the researchers who contributed to the TCGA data. We thank the researchers who built and maintained the online portals used in this work: cBio Portal, Firebrowse (Broad Institute), CGGA, and GEPIA.

## Authors contributions

G. M. and V.F. screened and took responsibility for the integrity and accuracy of the data analysis, performed the bioinformatic analysis, and wrote and edited the manuscript.

## Notes

### Competing Interest Statement

The authors have declared no competing interest.

### Funding Statement

This study did not receive any funding

### Summary of Updates

This revision reflects better the WHO glioma Classification from 2021 (CNS5)

